# Massive Open Online Course (MOOC) Opportunities on Health Education (HE) during of mandatory social isolation context

**DOI:** 10.1101/2021.03.01.21252702

**Authors:** Gandy Dolores-Maldonado, Jorge L. Cañari-Casaño, Rosalia Montero-Romainville, Germán Málaga

## Abstract

**Background:** Routine care for prevention and health promotion has reduced significantly due to the Covid-19 pandemic and mandatory social isolation measures. In this context, it is necessary to identify and describe Massive Open Online Courses (MOOCs) that provide opportunities for health education, promotion, and prevention aimed at the general population. The study is a systematic review of MOOCs on health education, health promotion, and prevention for the general population in a pandemic context.

**Methods:** We developed a search for MOOC courses aimed at the general population on health education, health promotion, and prevention in different MOOC available platforms. We executed a descriptive analysis of the main characteristics of the selected MOOCs.

**Results:** There were 117 MOOCs chosen on Health education, promotion, and prevention for the general population. Coursera (40.3%) was the platform that offered the highest quantity of MOOCs; more than half of MOOCs language was in English (52.9%). The median duration time in hours of the selected MOOCs was 11 (IR 6-15). The predominant themes were “Health promotion” (43%) and “Food and nutrition” (31%), and the origin was mainly from Europe (37.8%).

**Conclusions:** MOOCs offerings in Health Education (HE) is diverse, predominantly in English, of European origin, and in health promotion issues. This study opens an opportunity to multiply initiatives in different territories, considering other languages and topics more akin to each territorial reality, allowing it to be a more equitable learning opportunity in times of pandemic and compulsory social isolation.

**FUNDING:** None.

## INTRODUCTION

In December 2019, the COVID-19 pandemic was triggered by severe acute respiratory syndrome coronavirus 2 (SARS-Cov-2), revealing the fragility of the health systems of developing countries, to date March 1, in 2021, the cases amounted were 114,391,317 and 2,536,905 deaths worldwide (1) and the countries with the highest fatality due to covid-19 are mostly low-middle-income countries (LMIC) with precarious health systems or that already collapsed (2)(3).

The primary care services efforts have concentrated on containing the COVID-19 pandemic, for which there is a great concern about what could be neglected (4), reducing in access to medical doctors, drugs and growth monitoring during the lockdown period. (5) Also, disruptions in drug supply chains are likely associated with defaulters on immunization schedules, which may lead to future outbreaks of preventable diseases such as diphtheria. (6) Have been estimated that maternal and child neglect in LMIC could be devastating in a context where maternal deaths could increase up to 60% and infant mortality up to 41% (7). The control of endemic infectious diseases such as malaria (8), as well as chronic non-communicable diseases (NCDs) such as hypertension and diabetes, have been neglected or suspended (9), an increase in mental illnesses such as anxiety, depression, and suicide (10). Furthermore, there is a concern of the population to visit health systems for their routine care for fear of contagion (11). Against this, some countries implemented remote health care systems (teleconsultation) (12) and health communication campaigns. However, these strategies have been insufficient to cover the demand for health care affected by the pandemic and mandatory social isolation measures.

The pandemic context requires changes and identification of strategies that can help meet the indirect effects of neglect of diseases not related to Covid-19 in health systems. In this scenario, MOOCs could be an educational option to disseminate systematized courses on education, health promotion, and prevention aimed at the general population.

MOOCs, in the past, have been an opportunity for health education for developing countries. Among its advantages is global accessibility, flexible hours, multiple teaching tools, and generally free. MOOCs have been an educational response to emerging and re-emerging disease epidemics (13). However, to access these resources, inequities happened for developing countries, such as language barriers and technological access (14).

## METHODS

The study has conducted a digital search on MOOC platforms like Coursera, edX, FutureLearn, XuentangX, Udacity, Miríadax, Alison, Canvas Network, OpenWHO, among others to identify MOOCs with content related to health education (education, promotion, and prevention of health) aimed at the general public. Also, the search involved explored topics related to health, well-being, and medicine. We included terms as nutrition, healthy life, physical activity, medical care, healthy nutrition, mental health, and variants.

Three authors conducted the MOOC search manually and independently on the mentioned virtual platforms. The search development was in the month of June and December 2020. Likewise, we had to consult websites on larger platforms available in the world like Class Central (15) and MOOC List. We started the search of each virtual platform and examined MOOC contents with the terms described above. The eligibility criteria for selecting the MOOCs were that it evidences content related to health education, aimed at the general population; further, we consider being available for registration/access at the time of the search.

Subsequently, through a peer review, the researchers excluded MOOCs that showed highly specialized content or requested a prerequisite. MOOCs aimed at professionals or indicated that they were MOOCs for professional certification were also not considered, as well, only available for paid content. If a conflict or inconsistency existed about our exclusion criteria was solved through the deliberation peer review. We organized MOOCs by groups according to similar topics for a better description.

The data analysis was about the place of origin, principal language, and duration of the course. We used frequency measures to describe the categorical characteristics and dispersion measures to describe the hours. The analysis was using STATA version 16.

## RESULTS

With the established search criteria, a total of 217 MOOC courses were found on the different platforms, after excluding MOOC courses because they were specialized, unavailable, aimed at other target audiences, or without relation to health education. We selected 117 of the total MOOCs to be analyzed (Figure 1).

**Figure 1.**
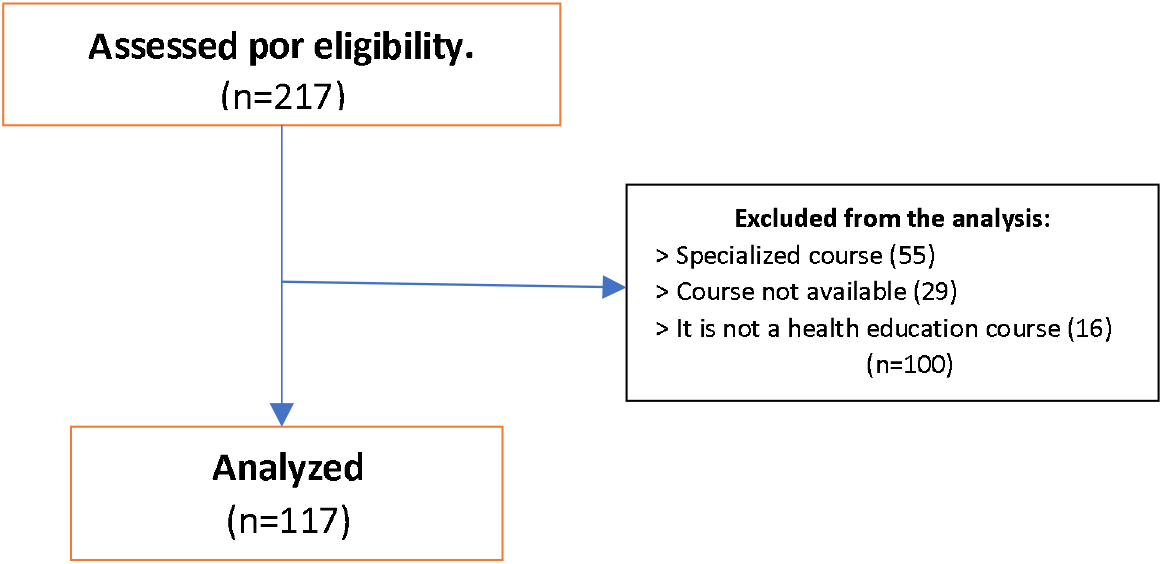
Flow diagram.

The 117 MOOCs analyzed were classified into six groups according to their content, with themes as health promotion, food and nutrition. more than 60% of the total MOOCs included. Regarding the duration of time (hours-the MOOCs found a median of 11 (RI 6-15) hours and the topics health promotion and community health / social right presented higher medians, as well as a minimum of 8.5 hours and a maximum of 16.5 hours (Table 1).

**Table 1.** Frequency, median and interquartile range of MOOC duration time.

| N° | Topics | N (%) | Median (RI) |
| --- | --- | --- | --- |
| 1 | Health promotion | 42 (35.9) | 12 (8.5-16.5) |
| 2 | Food and nutrition | 31 (26.5) | 9 (6-12) |
| 3 | Psychology and mental health | 23 (19.7) | 11 (5-16) |
| 4 | Health care | 13 (11.1) | 10 (6-12) |
| 5 | Climate change and health | 4 (3.4) | 2 (2-5) |
| 6 | Community health/ social rights Total | 4 (3.4) | 15 (13.5-16) |
|  | Total | 117 (100) | 11 (6-15) |

RI: Interquartile range

On Coursera were offered 1 (50%) and 12 (52.17%) from a total number of courses on health promotion and psychology and Mental Health topics, respectively. Likewise, 10 (32.26%) and 5 (38.46%) of total on food and nutrition and health care topics were offered on the Future Learn and EdX platforms, respectively (Table 2). Also, specify that all the courses around COVID19 (5 MOOCs were classified 4 in psychology and mental health and 1 in health promotion).

**Table 2.**
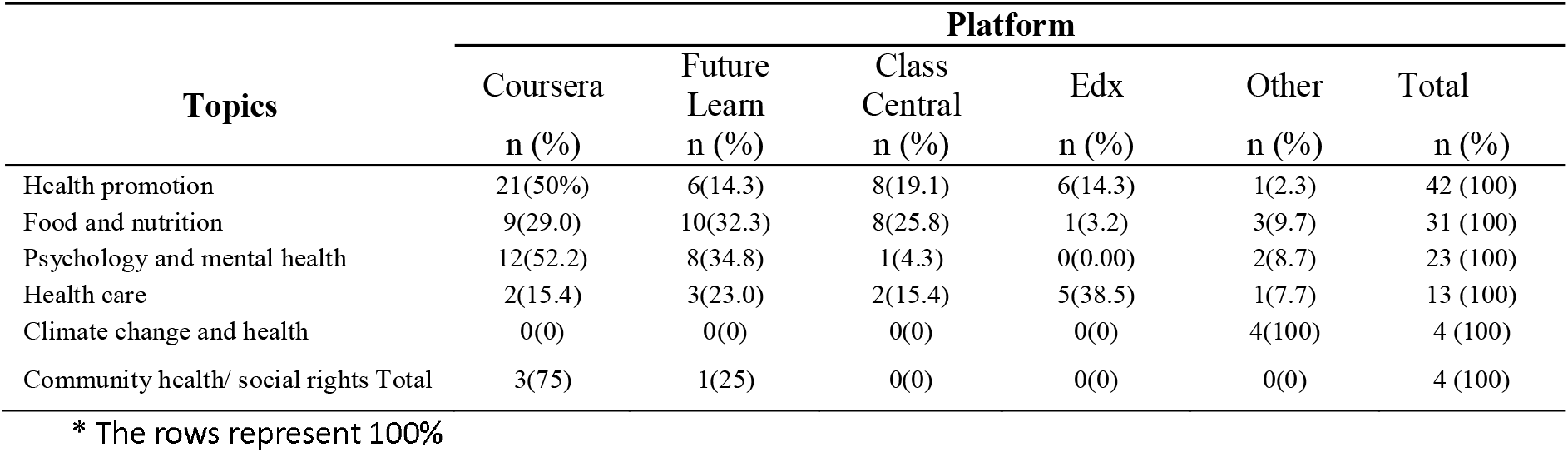
Distribution of MOOC topics by platform.

Regarding the principal language, was identified 62 (53%) of the MOOCs presented English as the language of preference, followed by Spanish 18 (15.4%) MOOC courses (data not shown). Likewise, the principal language on all topics was in the English language, with climate change and health topic exception (See Table 3).

**Table 3.**
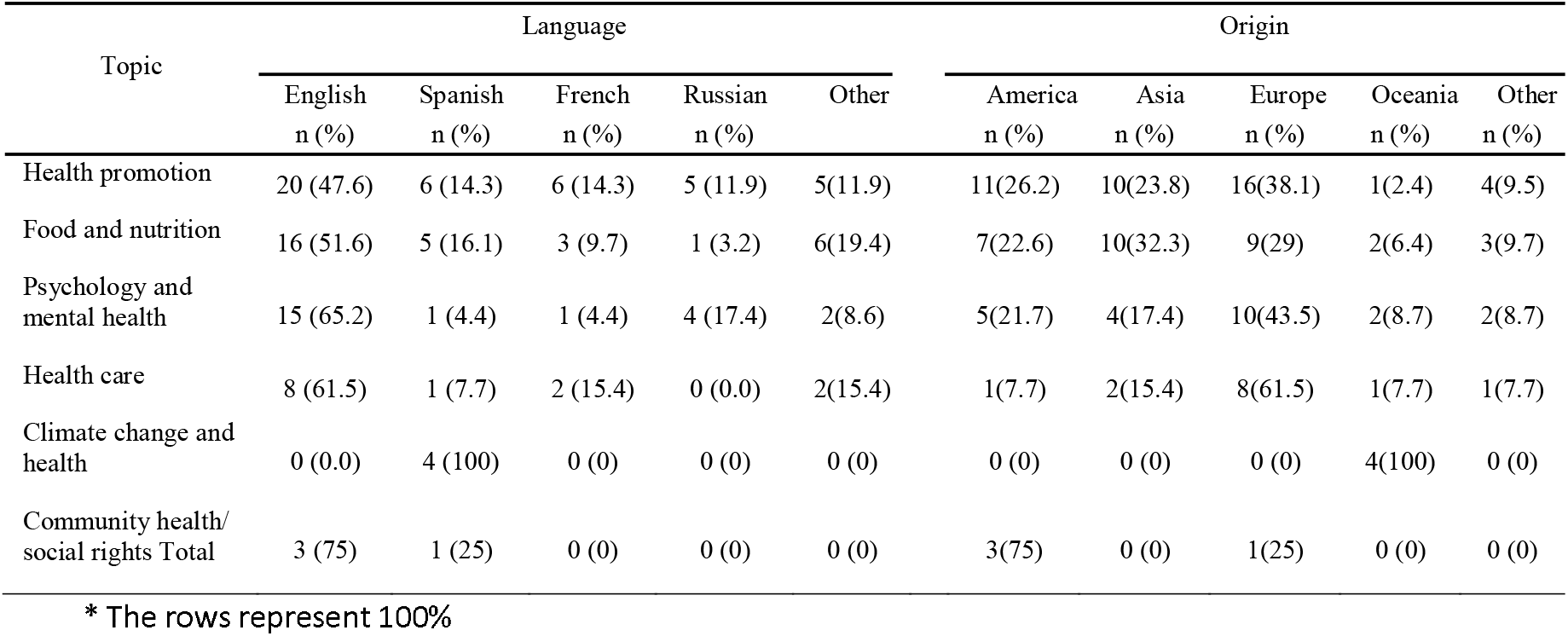
Distribution of MOOC topics by language and origin.

Regarding the origin of the MOOC courses, found that 44 (37.6) of the MOOCs were from Institutions in Europe, followed by America 27 (23.1) and Asia 26 (22.2) (Data not shown). In the case of America, 25 (21.4%) were from North America and 2 (1.7%) from South America (data not shown). The topics health promotion, psychology, and mental health and health care came mainly from universities in Europe; and the “Food and Nutrition” topic from Asia (See Table 3).

## DISCUSSION

We identified 117 MOOC courses on health promotion the majority are offered in English and are carried out mainly by institutions on the European continent and mostly found topics were health promotion and food and nutrition.

From the preliminary search, was evidence that a large part of the MOOCs maintained among their characteristics, possessing technical content of some specialty, learning units aimed at professionals (16)(17) others offering professional certification (13)(18)(19)(20), and some are not available upon request upon payment. Although we have not included these MOOCs in the study, it is essential to notice that it can be an indicator of the limited supply of MOOCs with a profile aimed at the general public or users of primary level care centers, with the content of free health education and aimed at prevention and health care. Consider this is extremely important for access to health education in a context of compulsory social isolation.

Regarding the predominant themes of health promotion and food and nutrition certain similarity was found with another study whose main topics were food, nutrition your health, and introduction to health nursing, courses were aimed at professionals (21). The MOOCs on health and medicine allow patients to acquire health education on specialized topics. They can understand disease implications, conditions, techniques, and available interventions around their disease, especially in the early stages. Besides, they can be useful some topics are still taboo, such as contraception, drug addiction, and acquired immunodeficiency syndrome (AIDS); courses focused on these topics help people educate themselves without having to visit an office.

The MOOCs found around psychology and mental health turn out to be a learning opportunity for stress management in times of compulsory social isolation. The results end up being part of recommendations to review said web-based interventions in mental health literacy promotion (22). It is because adolescents and young people are the ones who present more difficulties (23) for decision-making in health since when they search for information on the web (24), it is evident that they do not differentiate reliable information from the less reliable and that they do not know how to translate it into healthy behaviors (25).

Among other issues, community health and social rights take a position in the context of compulsory social isolation since many decisions about health can be taken collectively in the community environment (26), addition to many of them can be taken at the family level or by the influence of peers, without considering the repercussions of community leadership in some scenarios (27). Therefore, individual decisions can be even more relevant; For example, vaccination can affect a significant group of the population and have an impact on a higher incidence of some pathologies at the community level (28), especially when there is a population increase that declares not to be vaccinated even in front of the COVID19 pandemic (29).

Therefore, access to information through a MOOC could empower people who would not otherwise know about the options offered(21). This study shows that various institutions and organizations worldwide have seen MOOCs as an educational opportunity due to their relatively low cost (30) and whose success depends on the quality of their contents (31), the teacher’s strategies, and the focused courses.

Similar to previous studies was evidence that the Coursera platform was the one that hosted the largest number of MOOCs (21)(32). Regarding the origin of the MOOCs, the largest number were from developed countries (33), from institutions in Europe and North America, similar results were described in other studies (13)(21)(34), being, by default smaller quantity offered by Latin American countries (35). This predominant origin could be because more than half of the MOOCs were offered and developed in English (21)(13)(36) and only 22% in Spanish. Proof of this is that of the 23, 13, and 4 MOOCs on psychology and mental health, health care, community health / social rights, respectively, only one MOOC for each topic was in Spanish.

The majority offer of MOOCs in English may be a limitation for access and learning opportunities in times of pandemic for the Latin American population; as well as language, aspects such as the absence of a computer and internet, educational level (37) is also limited for access to MOOCs in times of compulsory social isolation.

Considering these courses were offered by developed countries as a fact. This issue could limit the topics addressed from being oriented to a different health reality from that of developing countries, where diseases such as anemia, malnutrition, or infectious diseases are the most frequent. This scenario could explain the high level of results from North America and Europe (38)(39) compared to South America, Africa, and Oceania (40) in MOOC courses. It was known courses were build based on a context and socioeconomic condition for a target population, and participation levels were higher when considered these variables (31). MOOCs with an approach based on the reality of LMIC (41) could be an opportunity, addressing issues such as chronic malnutrition, anemia, among frequent health problems that this population suffers, even more so in times of pandemic due to the restricted care of primary health centers to provide services on these issues.

Among the limitations were: The chosen courses are based exclusively on the authors’ criteria. The possibility of including studies that did not meet the inclusion criteria was lowered by performing the peer review. Courses classified into topics related to health education considered when compiling MOOCs for the review. However, if a MOOC has an incorrect classification, it would not have been identified for review. In cases MOOCs have been offered in languages different than English, we used Google Translate for content translation. Finally, the study aimed not to evaluate the quality of the contents in the MOOCs; however, almost all the MOOCs declared their institutional origin, which was predominantly universities.

Finally, the study showed most of the MOOC courses in health education aimed at the general population or users of health systems were framed mainly in the themes of health promotion and food and nutrition, originating from European institutions and North America and with a higher predominance of the English language.

MOOCs are showed as key tools to empower people, so in a pandemic context, the need to invest in alternative methods of dissemination of knowledge for knowledge-based empowerment would arise, covering the capacities of the general public that at present it is affected by not having access to care in health services of the first level of care. In addition to this, a critical shortage of human resources in health and healthcare, comprising a limited number of medical professors and limitations in physical infrastructures are reasons that increase the need to access online courses in health education of the level primary. Although the MOOCs’ origin was mainly from university institutions a future analysis of the quality of the contents must be addressed for greater comprehensiveness.

## Data Availability

Requests should be made to the correspondent author.

## AUTHORSHIP CONTRIBUTIONS

GDM participated in the conception and design of the article, data extraction, interpretation of the results and final writing. JCC participated in data extraction, statistical analysis, interpretation of results, and writing of the original draft. RMR participated in the data extraction and interpretation of results, the writing of the original draft, as well as in the preliminary translation of the manuscript. GMR participated in the interpretation of data through critical review and final writing.

## DECLARATION OF INTERESTS

We declare no competing interests.

## DATA SHARING

Requests should be made to Gandy Dolores-Maldonado

## ACKNOWLEDGMENTS

We express our acknowledgement to Corali Torres Paz de Fetaya for her review of the English version of our manuscript.

## Notes

### Competing Interest Statement

The authors have declared no competing interest.

### Author Declarations

The data used are freely accessible, therefore no informed consent was requested.

